# Effects of sex hormones on brain GABA and glutamate levels in a cis- and transgender cohort

**DOI:** 10.1101/2021.10.20.21265242

**Authors:** B Spurny-Dworak, P Handschuh, M Spies, U Kaufmann, R Seiger, M Klöbl, ME Konadu, MB Reed, V Ritter, P Baldinger-Melich, W Bogner, GS Kranz, R Lanzenberger

## Abstract

Sex hormones affect the GABAergic and glutamatergic neurotransmitter system as demonstrated in animal studies. However, human research has mostly been correlational in nature. Here, we aimed at substantiating causal interpretations of the interaction between sex hormones and neurotransmitter function by using magnetic resonance spectroscopy imaging (MRSI) to study the effect of gender-affirming hormone treatment (GHT) in transgender individuals.

Fifteen trans men (TM) with a DSM-5 diagnosis of gender dysphoria, and 15 age-matched cisgender women (CW) underwent MRSI before and after at least 12 weeks of GHT. Additionally, sex differences in neurotransmitter levels were evaluated in an independent sample of 80 cisgender men and 79 cisgender women. Mean GABA+ (a combination of GABA and macromolecules) and Glx (a combination of glutamate and glutamine) ratios to total creatine (GABA+/tCr, Glx/tCr) were calculated in five predefined regions-of-interest (hippocampus, insula, pallidum, putamen and thalamus).

Linear mixed models analysis revealed a significant measurement by gender identity effect (p_corr._ = 0.048) for GABA+/tCr ratios in the hippocampus, with the TM cohort showing decreased GABA+/tCr levels after GHT compared to CW. Moreover, analysis of covariance showed a significant sex difference in insula GABA+/tCr ratios (p_corr._ = 0.049), indicating elevated GABA levels in cisgender women compared to cisgender men.

Our study demonstrates GHT treatment-induced GABA+/tCr reductions in the hippocampus, indicating hormone receptor activation on GABAergic cells and testosterone-induced neuroplastic processes within the hippocampus. Moreover, elevated GABA levels in the female compared to the male insula highlight the importance of including sex as factor in future MRS studies.

## 1. Introduction

Sex hormones have widespread effects on the human brain. Several magnetic resonance spectroscopy (MRS) studies have attempted to investigate sex hormonal effects on a neurochemical level indirectly by studying sex differences or by examining metabolite levels across the menstrual cycle. For example, Grachev and colleagues found an effect of sex on various metabolites across brain regions (Grachev and Apkarian, 2000). Other studies observed sex differences in N-acetylaspartate (NAA) levels and choline/NAA (Wilkinson, 1997; Braun, 2002; Maudsley, 2009; Collet, 2021), while some studies reported no gender differences in NAA (Komoroski, 1999; Nagae-Poetscher, 2004; Jung, 2005). Especially neurotransmitter levels, regulating brain function, are of high interest. A study by Hadel et. al revealed sex-related differences in hippocampal glutamate levels (Hadel, 2013) while others observed higher levels of GABA, glutamate and a combined measure of glutamate and glutamine (Glx) in males compared to females in the dorsolateral prefrontal cortex (dlPFC) utilizing single-voxel MRS (O’Gorman, 2011). As concisely reviewed by Dubol and colleagues, a significant influence of sex hormones on brain functioning could be investigated throughout the menstrual cycle, suggesting a modulatory effect of fluctuating ovarian hormones especially on cortico-limbic brain regions (Dubol, 2021). Furthermore, sex hormones were shown to modulate brain structure and function in terms of activation and inhibition measured by structural and functional MRI along the menstrual cycle (Syan, 2017; Pletzer, 2018). Finally, Epperson and colleagues reported an influence of sex hormones during the menstrual cycle on cortical GABA levels in both smoking and non-smoking women (Epperson, 2002; Epperson, 2005).

In contrast to correlational research in humans, several animal treatment studies convincingly demonstrated sex hormone effects on metabolite levels. For example, estrogen treatment of male rats showed increases of glutamate, glutamine or Glx in the hippocampus, hypothalamus, frontal and parietal regions as well as increases in frontal GABA concentrations (Gomez, 2020). Moreover, an influence of androgen receptor activation on hippocampal glutamate response (Foradori, 2007) or antagonistic effects of glutamate on androgen receptors could be demonstrated (Shannon, 2019; Chiang and Park, 2020).

However, insights into metabolic adaptions upon interventional sex hormone applications in humans are scare. Here, transgender individuals undergoing gender-affirming hormone treatment (GHT) offer a unique investigatory framework to study the effect of high dosages of sex hormones, and to disentangle the impact of sex hormones from genetic sex and gender identity (for a review, see (Kranz, 2020)). In transgender individuals diagnosed with gender dysphoria (GD), the social gender does not match the biological sex assigned at birth. Hence, selected individuals with GD aim to undergo GHT in order to align the secondary sexual characteristics with their gender identity. Endocrine feminization of transfemales (TF; individuals with male sex assigned at birth but experiencing female gender identity) is accomplished via estrogen and progesterone treatment, whereas masculinization of transmales (TM; individuals with female sex assigned at birth but experiencing male gender identity) is done via testosterone treatment. A recent MRS study with a single-voxel positioned in the amygdala and hippocampus revealed differences of Cho/Cr and Glx/Cr ratios, as well as myo-inositol and glycine in the parietal cortex in transmale compared to cisgender male participants (Collet, 2021). However, to the best of our knowledge, no study has investigated the effects of sex hormones on a neurochemical level in people with GD undergoing GHT.

Hence, this study aimed to investigate the effects of GHT in TM participants compared to a cisgender women cohort (CW) using a 3D multi-voxel GABA-edited MRSI sequence. In addition, we collected baseline data of a large independent sample of 159 cisgender individuals (80 men, 79 women) in order to substantiate sex differences in neurotransmitter levels observed in previous underpowered studies.

## 2. Methods

### 2.1. Study design

This study is part of a larger longitudinal, case-control experiment (clinicaltrials.gov registration no. NCT02715232). Transgender individuals as well as age-matched cis controls were assessed before and after at least 12 weeks into GHT, see (Kranz, 2021). The current study focusses on the assessment of MRS data. Here, TM participants underwent MRS at baseline (M1), as well as after 12 weeks into testosterone treatment (M2) (for details see section 2.3). Plasma levels of estradiol, progesterone and testosterone were collected at each measurement. For comparison, cisgender women (CW) underwent two scans with the same time interval as TM subjects between M1 and M2.

### 2.2. Participants

Fifteen hormone-naïve TM participants (median+/-IQR age: 21 ± 7.5 years), with a DSM-5 diagnosis of GD and fifteen age-matched (± 3 years) cisgender women (median ± IQR age: 23 ± 4.5 years) were included in the analysis. In addition, 159 cisgender participants (80 men, mean age ± SD: 25.5 ± 5.3 years and 79 women, mean age ± SD: 25.4 ± 5.3 years) were assessed using MRS once. Baseline measurements of different studies (FWF: KLI 516, KLI 504 and NARSAD: 23741) were conducted between October 2017 and October 2020 using the same GABA-edited 3D MEGA-LASER MRSI sequence described in section 2.4. In individuals suffering from GD, the DSM-5 diagnosis was secured by means of psychological, psychiatric and psychotherapeutic expert opinions as well as by a clinical interview performed by a trained psychiatrist at the study site. All participants included in the analysis were free from any major internal or neurological disorder, psychiatric comorbidity (transgender participants) or psychiatric diagnosis (cisgender participants). General health was assessed based on medical history, physical examination, routine laboratory parameters and electrocardiogram. Psychiatric comorbidities (transgender subjects) or diagnoses (cisgender subjects) were excluded by a trained psychiatrist utilizing the Structured Clinical Interview for DSM-5. Exclusion criteria comprised substance abuse, MRI contraindications, pregnancy, current breastfeeding, as well as hormonal treatment in CW. Urine drug tests and pregnancy tests were performed at screening and before each MRI measurement. All participants provided written informed consent and received financial reimbursement. Baseline data from the 159 cisgender participants was pooled from different study protocols, all approved by the Ethics Committee of the Medical University of Vienna and conducted according to the Declaration of Helsinki.

### 2.3. Gender-affirming hormone therapy

TM participants received either 1000mg testosterone intramuscularly every 8-12 weeks (Nebido®) or alternatively 37.5-50 mg testosterone cream or gel transdermally per day (Testavan®). In some cases TM additionally received 75 μg desogestrel (Moniq Gynial®) per day or a triptorelin depot (Decapeptyl®) every 4-5 weeks (for details, see **supplementary table 1**). Due to the different treatment regimes, blood plasma levels of progesterone (ng/ml), estradiol (pg/ml) and testosterone (ng/ml) were quantified at each measurement and included as covariates in the statistical analyses. Missing hormone data was estimated using trimmed-scores regression with the standard settings utilizing the Missing Data Imputation v2 toolbox in MATLAB (Folch-Fortuny, 2015, 2016). The number of components with the minimum predicted residual sum of squares was retained. Imputation of missing hormone levels (CW: 3.3%, TM: 15.6%) was performed on log-transformed values due to the data distribution for each group separately treating each time point as different variable. Thresholds were sets at 1.5 times the interquartile range for back-transformed values to remove strong outliers.

### 2.4. MRSI measurements

All MRI measurements were performed on a 3 Tesla MR Magnetom Prisma system (Siemens Medical, Erlangen, Germany) installed at the High-field MR Center, Department of Biomedical Imaging and Image-guided Therapy, Medical University of Vienna, using a 64-channel head coil. For accurate volume of interest (VOI) placement and mask extraction for region-of-interest (ROI)-based quantification we acquired T1-weighted images (TE = 1800 ms, TR = 2.37 ms, 208 slices, 288 × 288 matrix size, voxel size 1.15 × 1.15 × 0.85 mm) prior to each MRSI scan.

For spectroscopic measurements a spiral-encoded 3D GABA-edited MEGA-LASER MRSI sequence with an echo time of 68 ms described in (Bogner, 2014a) was used. Real-time correction for rigid-body motion bias and center frequency was applied (Bogner, 2014b) using volumetric, dual contrast, echo planar imaging-based navigators with a repetition time of 1.6 s, updated every 3.2 s. The VOI was placed parallel to the anterior commissure–posterior commissure line to cover the hippocampus and insula bilaterally, with a VOI = 80 (l-r) × 90 (a-p) × 80 (s-i) mm^3^ and a field of view (FOV) = 160 × 160 × 160mm^3^ (see **Figure 1**). The acquired matrix size of 10 × 10 × 10 (i.e., ∼4 cm3 nominal voxel size) was interpolated to a 16 × 16 × 16 matrix (i.e., ∼1 cm^3^ nominal voxel size) during spectral processing steps. MEGA-editing pulses utilizing 60 Hz Gaussian pulses of 14.8 ms duration were set to 1.9 ppm during EDIT-ON acquisition. 32 acquisition-weighted averages and two-step phase cycling were employed, resulting in a total scan time of 15:09 min. Advanced Siemens shimming procedure with manual adjustments was conducted prior to each MRSI scan.

**Figure 1:**
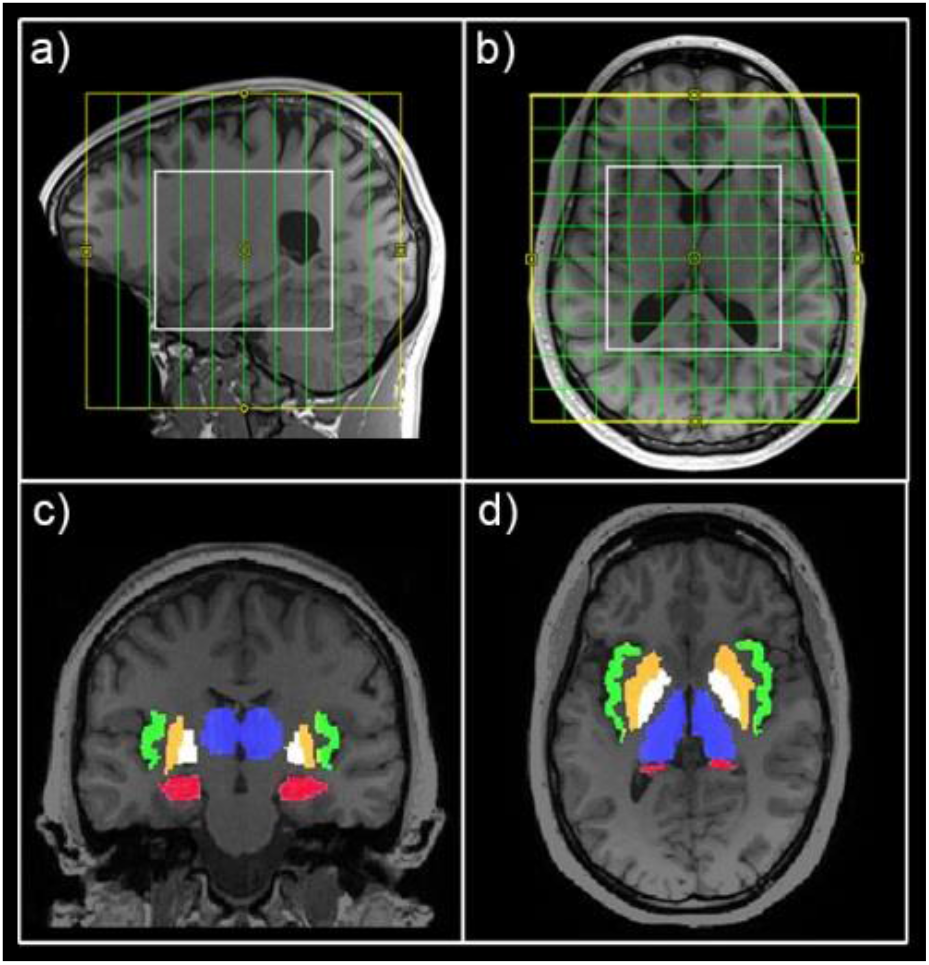
Field of view position and regions of interest. Field of view (yellow) and volume of interest (white) are shown in defaced sagittal (a) and horizontal (b) view. Exemplary masks for the hippocampus (red), insula (green), putamen (orange), pallidum (white) and thalamus (blue) are presented in coronal (c) and horizontal (d) view.

### 2.5. MRSI data analysis

A combination of MATLAB (R2013a, MathWorks, Natick, MA, USA), Bash (4.2.25, Free Software Foundation, Boston, MA, USA), MINC (2.0, MINC Tools, McConnell Brain Imaging Center, Montreal, QC, Canada) and LCModel software (6.3–1, S. Provencher, LCModel, Oakville, ON, Canada) was used for the quantification of all spectra inside the VOI. The GAMMA library was used for the creation of two different basis sets, one for the non-edited spectra (containing 21 brain metabolites, including total creatine (tCr)) and one for the difference spectrum (containing GABA+ and Glx among others) (Hnilicova, 2016). Exemplary spectra can be seen in **supplementary figure 1**. Cramér–Rao lower bounds (CRLB) thresholds were set at 30% and spectra were visually inspected. An ROI-based quantification (described in (Spurny, 2019)) was applied for the analysis of GABA+ (a combination of GABA and macromolecules) and Glx (glutamate+glutamine) ratios to total creatine (GABA+/tCr and Glx/tCr) in the hippocampus, insula, putamen, pallidum and thalamus. ROI selection was based on previous studies showing structural adaptions after GHT in trans cohorts (Spizzirri, 2018; Kranz, 2020). In short, FreeSurfer was used for automated segmentation of structural images and mask extraction, spectral maps of GABA+, Glx and tCr were interpolated to the resolution of structural images (288×288×208), overlaid with the derived masks and mean GABA+/tCr and Glx/tCr ratios calculated within each ROI. ROIs with < 90% valid voxels, due to CRLB thresholds, were excluded from the analyses.

### 2.6. Statistical analysis

To investigate the effects of GHT, we estimated linear mixed effects models (LMM) including measurement (M1 and M2) and group (TM, CW). For LMM analyses gender identity was included as factor and measurement as repeated factor, testing for fixed effects in gender identity, measurement and a gender identity x measurement interaction. Separate models were conducted for each ROI and neurotransmitter ratio. Model selection for the repeated covariance type was based on the minimal Akaike-information criterion. Age and plasma levels of progesterone, estradiol and testosterone were included as covariates in each model, whereas hormonal levels were mean-centered within each group and time point. To detect possible correlations between changes in hormone levels and neurotransmitter over time, a Spearman correlation analysis within each ROI, group and neurotransmitter ratio was performed. Finally, sex differences in neurotransmitter levels in the independent sample of 80 cisgender men and 79 cisgender women were evaluated using analyses of covariance (ANCOVA). Separate models were conducted for each ROI and neurotransmitter. Age was included as covariate in each model. Results within each analysis (LMM, Spearman correlation and ANCOVA) were corrected for multiple models using the Bonferroni method.

## 3. Results

### Effects of GHT

Mean time between measurements was 146±30 days for TM individuals and 166±40 days for CW subjects. Number of data points within each ROI that passed quality control is presented in **supplement table 2**. Linear mixed models for GABA+/tCr ratios in the hippocampus revealed a significant measurement by gender identity effect (p_corr.=_0.048; Bonferroni corrected for region (5), neurotransmitter ratio (2) and group (2)) with the TM cohort showing decreased hippocampal GABA+/tCr levels after GHT compared to the cisgender group (see **Figure 2**). No main or interaction effects could be shown for GABA+/tCr in other ROIs or for Glx/tCr ratios in any region.

**Figure 2:**
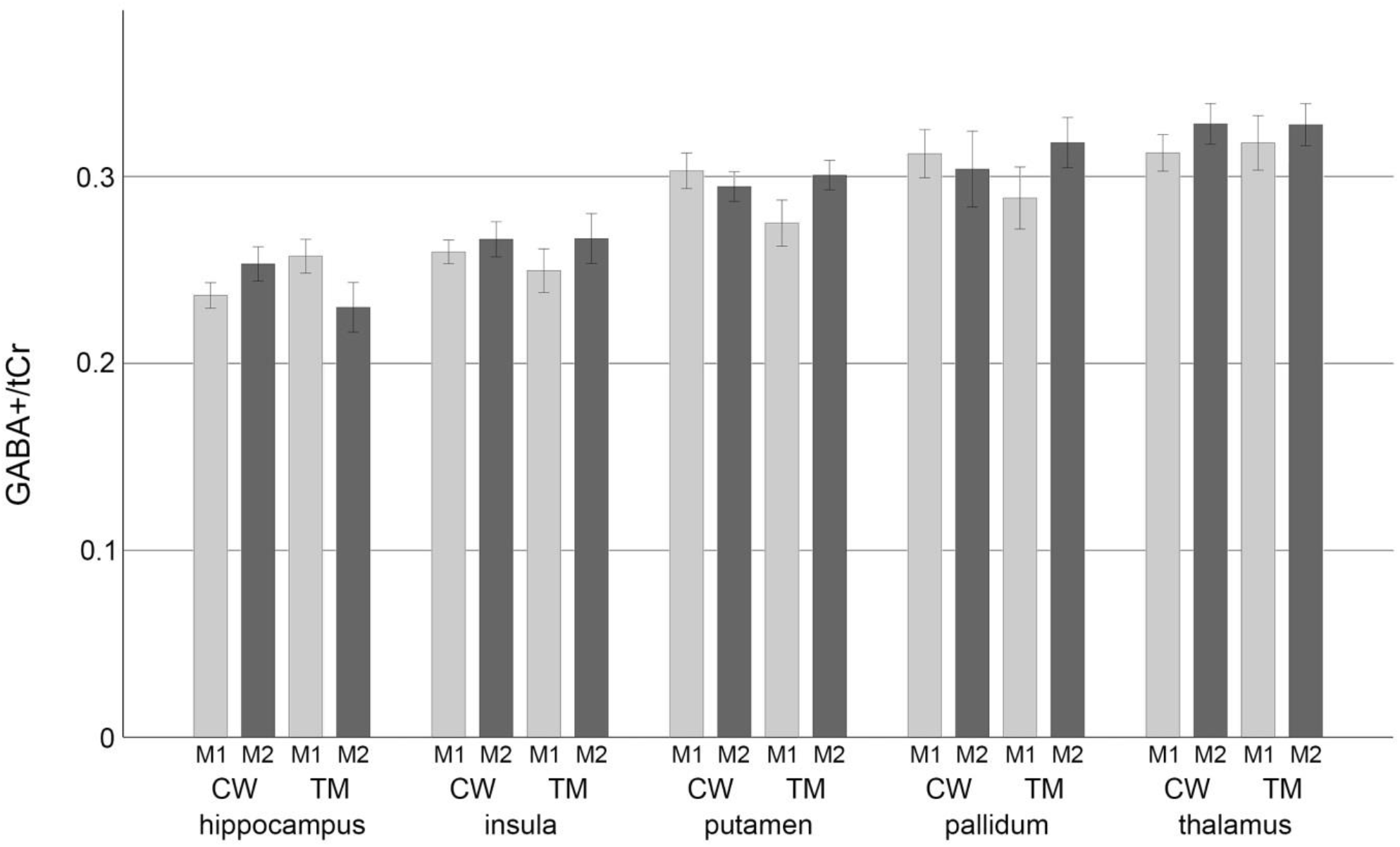
Bars denote mean GABA+/tCr ratios of cisgender women (=CW) and trans men (=TM) at M1 (light grey) and M2 (dark grey) within each region of interest. Brackets showing the standard error. GABA+= GABA + macromolecules; tCr= total creatine

### Relationship of neurotransmitter ratios and plasma hormone levels

No significant correlations between hormone levels of estradiol, progesterone or testosterone and neurotransmitter ratios could be detected within TM or CW in any ROI.

### Sex differences in neurotransmitter levels in cisgender male and female subjects

ANCOVAs revealed a significant difference in GABA+/tCr in the insula between cisgender men and cisgender women (p_corr._ = 0.049; Bonferroni corrected for 10 comparisons), showing higher levels in females compared to males. No significant differences in GABA+/tCr could be shown in any other ROI or Glx/tCr ratios across all regions investigated (see **Figure 3**). Mean metabolite concentrations within each ROI are shown in **Table 1**.

**Figure 3:**
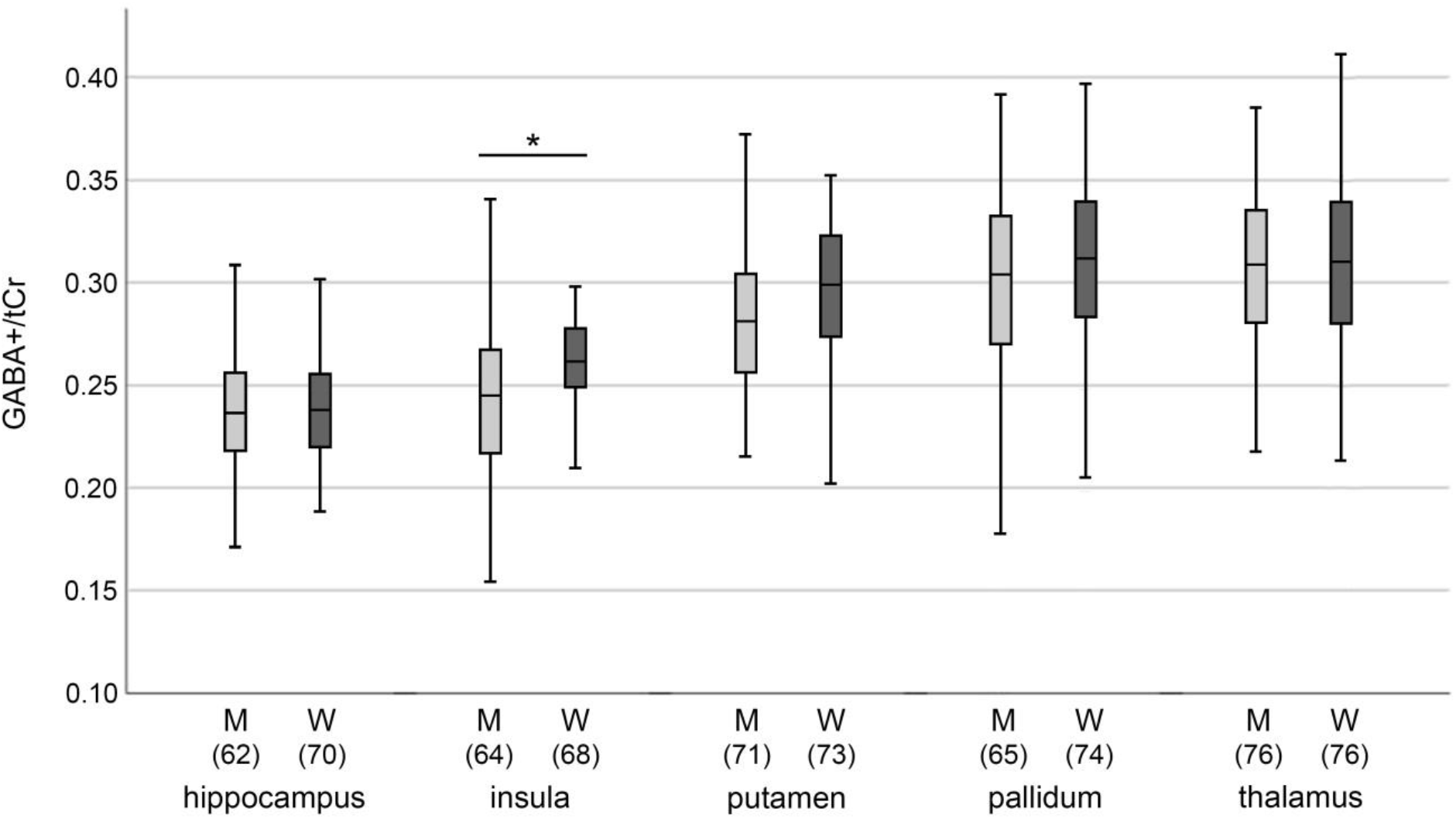
Mean baseline GABA+/tCr ratios of cisgender men (M) and cisgender women (W) subjects. Brackets represent number of participants within each group.

**Table 1:**
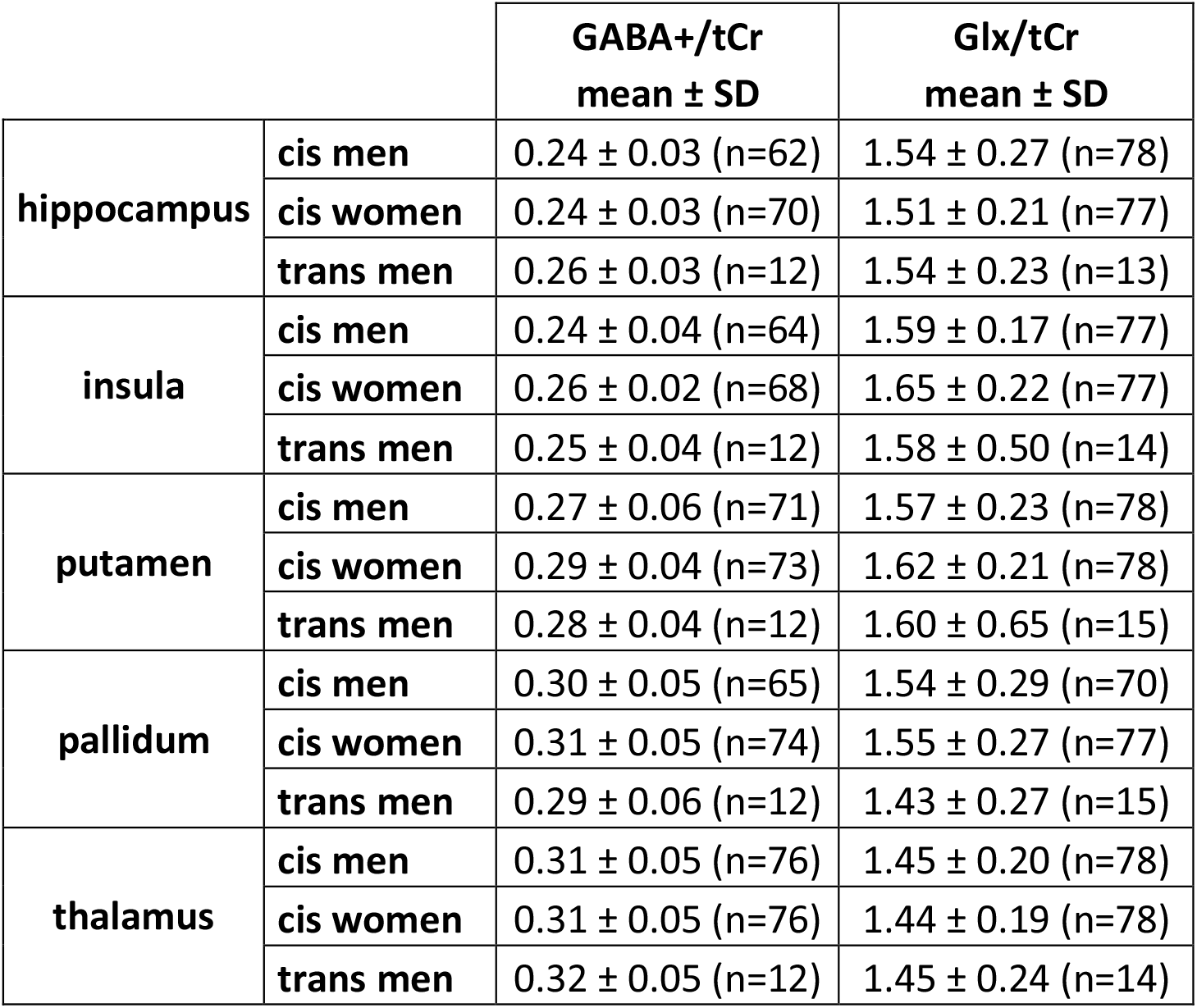
Mean metabolite concentrations of cisgender men, cisgender women and trans men at baseline

## 4. Discussion

Here, we describe a significant reduction of GABA+/tCr ratios in the hippocampus after gender-affirming hormone therapy in TM individuals compared to a female cisgender cohort. No effects on Glx/tCr ratios were found in other ROIs investigated. The correlation analysis performed did not show a relationship between changes in hormone levels and neurotransmitter ratios in any region. Nevertheless, a significant difference in GABA+/tCr levels in the insula between cisgender men and cisgender women at baseline was revealed in an independent sample.

Several studies showed an impact of androgens on hippocampal structure and function inducing complex responses. While these findings were primarily shown in animals, human studies were able to correlate (atypical) androgen levels with the incidence of neurodegenerative disorders or cognitive function (Driscoll and Resnick, 2007; Barron and Pike, 2012). In addition, two human studies investigating the effects of GHT in gender dysphoria showed no changes in hippocampal volumes after testosterone treatment in TM subjects (Seiger, 2016; Burke, 2018), whereas effects on white matter microstructure could be described within this region (Kranz, 2017).

While our study revealed no differences in the glutamatergic system we report effects of GHT on the hippocampal GABAergic system. The hippocampus is a brain region that is rich in hormone receptors (Milner, 2001; Tabori, 2005; Rai and Jeswar, 2012) and known for local hormone production in neurons and glia cells (Hojo, 2011). Moreover, this region shows properties for testosterone metabolism (Yague, 2010). Testosterone is converted to estradiol and dihydrotestosterone (DHT) in neurons and glia cells. In astrocytes, DHT in turn can be converted to 5α-androstane-3α,17β-diol, which shows modulatory properties of GABA on the GABA_A_ receptor (Reddy, 2004). Thus, GHT in TM subjects inevitably exposes different cell and tissue types to testosterone and its metabolites, all potentially affecting brain neurotransmitter levels. The interplay between sex hormones, brain structure and function as well as neurotransmitter systems was mainly described in animal studies so far. Receptors for androgens and estradiol can be found in cell nuclei, the plasma membrane or synaptic vesicles in rat hippocampal neurons (Kerr, 1995; Tabori, 2005). Testosterone treatment was shown to induce synaptic spine growth in the hippocampus of both male and ovariectomized female rats (Leranth, 2003; Leranth, 2004; MacLusky, 2006), while estrogens were reported to influence plasticity and spine formation via the N-methyl-D-aspartate (NMDA) receptor function (McEwen, 2012) and to modulate hippocampal neurogenesis (Galea, 2006; McEwen and Milner, 2017). Modulation of synaptic plasticity and spine formation of androgens is thought to be mediated via brain-derived neurotrophic factor (BDNF) in the mossy fiber system (Atwi, 2016). Androgens were reported to affect BDNF expression which in turn mediates androgen effects (Scharfman and MacLusky, 2014). Atwi and colleagues hypothesized that increased BDNF expression in the mossy fiber system could preferentially activate GABAergic interneurons and in consequence modulate spine formation and activity levels, since BDNF potentiates presynaptic GABA synthesis in interneurons, which are innervated by a huge amount of mossy fibers (Ohba, 2005; Atwi, 2016).

Here we report GABA reductions after testosterone treatment for the first time in human TM subjects with GD compared to cisgender women. GD is often accompanied by psychiatric disorders and increased rates in suicidality (Cole, 1997; Peterson, 2017). Especially major depressive disorder (MDD), characterized by altered neurotransmission and BDNF levels (Brambilla, 2003; Sanacora, 2012; Kishi, 2017), showed a high prevalence in individuals with GD (Heylens, 2014). Animal studies revealed a relationship between sex hormone receptors and the regulation of depressive-like behavior via the GABAergic system (Selakovic, 2019). While previous studies showed an impact of antidepressant treatment on hippocampal neurotransmitter levels (Silberbauer, 2020; Spurny, 2021), GHT follows similar patterns in mediating hippocampal neurotransmitter levels. Thus, GHT shows properties to influence hippocampal neuronal networks that were shown to be involved in MDD and other psychiatric diseases on a molecular level via hormone receptors and changes in BDNF levels. Interestingly, GABA levels in the hippocampus were shown to follow a similar pattern after treatment with the NMDA antagonist ketamine which is known to activate BDNF-mediated neuroplastic processes (Silberbauer, 2020). Ketamine is hypothesized to exert its antidepressant properties via NMDA activation on GABAergic interneurons as well as binding the tropomyosin receptor kinase B (TRKB) receptor and by activating BDNF-mediated neuroplastic processes via the mechanistic target of rapamycin complex (mTOR) pathway and α-amino-3-hydroxy-5-methyl-4-isoxazolepropionic acid (AMPA) activation (Autry, 2011; Duman, 2012; Zanos, 2016; Zanos and Gould, 2018; Casarotto, 2021). It can be assumed that effects of BDNF on GABAergic interneurons along with neuroplastic properties of androgens and estrogens are reflected in altered GABA+/tCr ratios similar to ketamine. Various mechanisms can potentially contribute to changes in GABA+/tCr ratios observed in this analysis. BDNF-mediated alterations in GABA synthesis (Atwi, 2016), effects of testosterone and its metabolites on hippocampal GABA concentrations via hormone receptors located GABAergic interneurons (Tabori, 2005; Rai and Jeswar, 2012) or interference with local hormone production (Hojo, 2011) reflected in altered neurotransmitter ratios are possible explanations that need to be clarified in future approaches.

In addition to the investigation of the effects of GHT in TM, we compared GABA+/tCr and Glx/tCr levels between cisgender men and women, measured once, in a big study cohort. Our analysis of 159 individuals revealed a significant difference in GABA+/tCr ratios in the insula. Hence, the sample size of our comparisons outnumbers most of the previous comparisons by far. Existing studies assessing gender effects on GABA levels via MRS have reported varying results and were restricted to a handful of brain regions. Sex differences in GABA ratios are not surprising, since GABA was shown to shape sex dimorphism in the developmental brain modulated by sex hormones (McCarthy, 2002). Moreover, studies revealed significant sex-dependent difference in α1 subunit expression of the GABA receptor in temporal cortical regions (Pandya, 2019) or differences between men and women in the GABAergic system following acute stress (Skilbeck, 2010). Single-voxel MRS studies either reported no sex differences (Hu, 2013) or revealed higher levels of GABA in male subjects, e.g., in the medial-parietal cortex (Saleh, 2017) or in the occipital cortex in health and MDD (Sanacora, 1999). Moreover, O’Gorman and colleagues reported higher levels of GABA and glutamate in the dlPFC in a relatively small cohort of cisgender men compared to women (O’Gorman, 2011). In contrast to their findings, we reported higher GABA levels in cisgender women compared to cisgender men in the insula. The insula is highly involved in body awareness, perception and ownership (Tsakiris, 2007; Craig, 2009). Sex differences in volume (Ruigrok, 2014), GABA_A_ subunit expression (Pandya, 2019) and activation patterns in body (self-)perception, as well as during sexual desire and arousal could be shown within this region (Poeppl, 2016; Burke, 2019). Thus, functional differences may be reflected in altered neurotransmitter concentrations. Especially alterations in hormonal levels of estradiol or progesterone during the menstrual cycle have been argued to cause changes in GABA concentrations in cisgender female subjects (Pandya, 2019). These fluctuations in GABA levels could be correlated with changes in mood (Backstrom, 2011). Although we were not able to show correlations of neurotransmitter ratios and hormone levels in TM subjects, hormone fluctuations during the menstrual cycle in cisgender women may explain sex-differences in neurotransmitter levels. Our findings in a profound sample size emphasize differences in neurotransmitter concentrations between genders and thereby highlight the importance for a correction for gender in statistical analyses of MRS studies especially for GABA ratios.

## Conclusion

This work describes the tight interplay between hormonal treatment and neurotransmitter systems in the human brain. The reported decrease in hippocampal GABA+/tCr ratios after testosterone treatment in TM participants compared to cisgender women may result from hormone-induced neuroplastic processes in combination with hormone receptor activation on interneurons that could be previously shown in animal studies. However, this is the first human study reporting GABA adaptions after GHT in TM subjects with gender dysphoria. Moreover, differences in baseline GABA+/tCr ratios in the insula could be reported in a cisgender cohort of 159 subjects adding evidence of sex differences in neurotransmitter levels reported in previous studies. Hence, a correction for sex in future MRSI analyses, especially when investigating GABA levels, is highly recommended.

## Data Availability

All data produced in the present study are available upon reasonable request to the authors

## Data availability statement

Due to data protection laws processed data is available from the authors upon reasonable request. Please contact with any questions or requests.

## References

Atwi S, McMahon D, Scharfman H, MacLusky NJ (2016) Androgen Modulation of Hippocampal Structure and Function. The Neuroscientist : a review journal bringing neurobiology, neurology and psychiatry 22:46–60, DOI: 10.1177/1073858414558065.

Autry AE, Adachi M, Nosyreva E, Na ES, Los MF, Cheng PF, Kavalali ET, Monteggia LM (2011) NMDA receptor blockade at rest triggers rapid behavioural antidepressant responses. Nature 475:91–95, DOI: 10.1038/nature10130.

Backstrom T, Haage D, Lofgren M, Johansson IM, Stromberg J, Nyberg S, Andreen L, Ossewaarde L, van Wingen GA, Turkmen S, Bengtsson SK (2011) Paradoxical effects of GABA-A modulators may explain sex steroid induced negative mood symptoms in some persons. Neuroscience 191:46–54, DOI: 10.1016/j.neuroscience.2011.03.061.

Barron AM, Pike CJ (2012) Sex hormones, aging, and Alzheimer’s disease. Frontiers in bioscience 4:976–997, DOI.

Bogner W, Gagoski B, Hess AT, Bhat H, Tisdall MD, van der Kouwe AJ, Strasser B, Marjanska M, Trattnig S, Grant E, Rosen B, Andronesi OC (2014a) 3D GABA imaging with real-time motion correction, shim update and reacquisition of adiabatic spiral MRSI. Neuroimage 103:290–302, DOI: 10.1016/j.neuroimage.2014.09.032.

Bogner W, Hess AT, Gagoski B, Tisdall MD, van der Kouwe AJ, Trattnig S, Rosen B, Andronesi OC (2014b) Real-time motion-and B0-correction for LASER-localized spiral-accelerated 3D-MRSI of the brain at 3T. Neuroimage 88:22–31, DOI: 10.1016/j.neuroimage.2013.09.034.

Brambilla P, Perez J, Barale F, Schettini G, Soares JC (2003) GABAergic dysfunction in mood disorders. Mol Psychiatry 8:721–737, 715, DOI: 10.1038/sj.mp.4001362.

Braun CM, Boulanger Y, Labelle M, Khiat A, Dumont M, Mailloux C (2002) Brain metabolic differences as a function of hemisphere, writing hand preference, and gender. Laterality 7:97–113, DOI: 10.1080/13576500143000212.

Burke SM, Majid DSA, Manzouri AH, Moody T, Feusner JD, Savic I (2019) Sex differences in own and other body perception. Hum Brain Mapp 40:474–488, DOI: 10.1002/hbm.24388.

Burke SM, Manzouri AH, Dhejne C, Bergstrom K, Arver S, Feusner JD, Savic-Berglund I (2018) Testosterone Effects on the Brain in Transgender Men. Cereb Cortex 28:1582–1596, DOI: 10.1093/cercor/bhx054.

Casarotto PC, Girych M, Fred SM, Kovaleva V, Moliner R, Enkavi G, Biojone C, Cannarozzo C, Sahu MP, Kaurinkoski K, Brunello CA, Steinzeig A, Winkel F, Patil S, Vestring S, Serchov T, Diniz C, Laukkanen L, Cardon I, Antila H, Rog T, Piepponen TP, Bramham CR, Normann C, Lauri SE, Saarma M, Vattulainen I, Castren E (2021) Antidepressant drugs act by directly binding to TRKB neurotrophin receptors. Cell 184:1299–1313 e1219, DOI: 10.1016/j.cell.2021.01.034.

Chiang VS, Park JH (2020) Glutamate in Male and Female Sexual Behavior: Receptors, Transporters, and Steroid Independence. Frontiers in behavioral neuroscience 14:589882, DOI: 10.3389/fnbeh.2020.589882.

Cole CM, O’Boyle M, Emory LE, Meyer WJ, 3rd (1997) Comorbidity of gender dysphoria and other major psychiatric diagnoses. Archives of sexual behavior 26:13–26, DOI: 10.1023/a:1024517302481.

Collet S, Bhaduri S, Kiyar M, T’Sjoen G, Mueller S, Guillamon A (2021) Characterization of the (1)H-MRS Metabolite Spectra in Transgender Men with Gender Dysphoria and Cisgender People. Journal of clinical medicine 10:DOI: 10.3390/jcm10122623.

Craig AD (2009) How do you feel--now? The anterior insula and human awareness. Nat Rev Neurosci 10:59–70, DOI: 10.1038/nrn2555.

Driscoll I, Resnick SM (2007) Testosterone and cognition in normal aging and Alzheimer’s disease: an update. Current Alzheimer research 4:33–45, DOI: 10.2174/156720507779939878.

Dubol M, Epperson CN, Sacher J, Pletzer B, Derntl B, Lanzenberger R, Sundstrom-Poromaa I, Comasco E (2021) Neuroimaging the menstrual cycle: A multimodal systematic review. Frontiers in neuroendocrinology 60:100878, DOI: 10.1016/j.yfrne.2020.100878.

Duman RS, Li N, Liu RJ, Duric V, Aghajanian G (2012) Signaling pathways underlying the rapid antidepressant actions of ketamine. Neuropharmacology 62:35–41, DOI: 10.1016/j.neuropharm.2011.08.044.

Epperson CN, Haga K, Mason GF, Sellers E, Gueorguieva R, Zhang W, Weiss E, Rothman DL, Krystal JH (2002) Cortical gamma-aminobutyric acid levels across the menstrual cycle in healthy women and those with premenstrual dysphoric disorder: a proton magnetic resonance spectroscopy study. Arch Gen Psychiatry 59:851–858, DOI: yoa10025 [pii].

Epperson CN, O’Malley S, Czarkowski KA, Gueorguieva R, Jatlow P, Sanacora G, Rothman DL, Krystal JH, Mason GF (2005) Sex, GABA, and nicotine: the impact of smoking on cortical GABA levels across the menstrual cycle as measured with proton magnetic resonance spectroscopy. Biol Psychiatry 57:44–48, DOI: 10.1016/j.biopsych.2004.09.021.

Folch-Fortuny A, Arteaga F, Ferrer A (2015) PCA model building with missing data: New proposals and a comparative study. Chemometrics and Intelligent Laboratory Systems 146:77–88, DOI: 10.1016/j.chemolab.2015.05.006.

Folch-Fortuny A, Arteaga F, Ferrer A (2016) Missing Data Imputation Toolbox for MATLAB. Chemometrics and Intelligent Laboratory Systems 154:93–100, DOI: 10.1016/j.chemolab.2016.03.019.

Foradori CD, Werner SB, Sandau US, Clapp TR, Handa RJ (2007) Activation of the androgen receptor alters the intracellular calcium response to glutamate in primary hippocampal neurons and modulates sarco/endoplasmic reticulum calcium ATPase 2 transcription. Neuroscience 149:155–164, DOI: 10.1016/j.neuroscience.2007.06.054.

Galea LA, Spritzer MD, Barker JM, Pawluski JL (2006) Gonadal hormone modulation of hippocampal neurogenesis in the adult. Hippocampus 16:225–232, DOI: 10.1002/hipo.20154.

Gomez A, Cerdan S, Perez-Laso C, Ortega E, Pasaro E, Fernandez R, Gomez-Gil E, Mora M, Marcos A, Del Cerro MCR, Guillamon A (2020) Effects of adult male rat feminization treatments on brain morphology and metabolomic profile. Hormones and behavior 125:104839, DOI: 10.1016/j.yhbeh.2020.104839.

Grachev ID, Apkarian AV (2000) Chemical heterogeneity of the living human brain: a proton MR spectroscopy study on the effects of sex, age, and brain region. Neuroimage 11:554–563, DOI: 10.1006/nimg.2000.0557.

Hadel S, Wirth C, Rapp M, Gallinat J, Schubert F (2013) Effects of age and sex on the concentrations of glutamate and glutamine in the human brain. J Magn Reson Imaging 38:1480–1487, DOI: 10.1002/jmri.24123.

Heylens G, Elaut E, Kreukels BP, Paap MC, Cerwenka S, Richter-Appelt H, Cohen-Kettenis PT, Haraldsen IR, De Cuypere G (2014) Psychiatric characteristics in transsexual individuals: multicentre study in four European countries. The British journal of psychiatry : the journal of mental science 204:151–156, DOI: 10.1192/bjp.bp.112.121954.

Hnilicova P, Povazan M, Strasser B, Andronesi OC, Gajdosik M, Dydak U, Ukropec J, Dobrota D, Trattnig S, Bogner W (2016) Spatial variability and reproducibility of GABA-edited MEGA-LASER 3D-MRSI in the brain at 3 T. NMR Biomed 29:1656–1665, DOI: 10.1002/nbm.3613.

Hojo Y, Higo S, Kawato S, Hatanaka Y, Ooishi Y, Murakami G, Ishii H, Komatsuzaki Y, Ogiue-Ikeda M, Mukai H, Kimoto T (2011) Hippocampal synthesis of sex steroids and corticosteroids: essential for modulation of synaptic plasticity. Frontiers in endocrinology 2:43, DOI: 10.3389/fendo.2011.00043.

Hu Y, Chen X, Gu H, Yang Y (2013) Resting-state glutamate and GABA concentrations predict task-induced deactivation in the default mode network. J Neurosci 33:18566–18573, DOI: 10.1523/JNEUROSCI.1973-13.2013.

Jung RE, Haier RJ, Yeo RA, Rowland LM, Petropoulos H, Levine AS, Sibbitt WL, Brooks WM (2005) Sex differences in N-acetylaspartate correlates of general intelligence: an 1H-MRS study of normal human brain. Neuroimage 26:965–972, DOI: 10.1016/j.neuroimage.2005.02.039.

Kerr JE, Allore RJ, Beck SG, Handa RJ (1995) Distribution and hormonal regulation of androgen receptor (AR) and AR messenger ribonucleic acid in the rat hippocampus. Endocrinology 136:3213–3221, DOI: 10.1210/endo.136.8.7628354.

Kishi T, Yoshimura R, Ikuta T, Iwata N (2017) Brain-Derived Neurotrophic Factor and Major Depressive Disorder: Evidence from Meta-Analyses. Frontiers in psychiatry 8:308, DOI: 10.3389/fpsyt.2017.00308.

Komoroski RA, Heimberg C, Cardwell D, Karson CN (1999) Effects of gender and region on proton MRS of normal human brain. Magn Reson Imaging 17:427–433, DOI: 10.1016/s0730-725x(98)00186-6.

Kranz GS, Seiger R, Kaufmann U, Hummer A, Hahn A, Ganger S, Tik M, Windischberger C, Kasper S, Lanzenberger R (2017) Effects of sex hormone treatment on white matter microstructure in individuals with gender dysphoria. Neuroimage 150:60–67, DOI: 10.1016/j.neuroimage.2017.02.027.

Kranz GS, Spies M, Vraka C, Kaufmann U, Klebermass EM, Handschuh PA, Ozenil M, Murgas M, Pichler V, Rischka L, Nics L, Konadu ME, Ibeschitz H, Traub-Weidinger T, Wadsak W, Hahn A, Hacker M, Lanzenberger R (2021) High-dose testosterone treatment reduces monoamine oxidase A levels in the human brain: A preliminary report. Psychoneuroendocrinology 133:105381, DOI: 10.1016/j.psyneuen.2021.105381.

Kranz GS, Zhang BBB, Handschuh P, Ritter V, Lanzenberger R (2020) Gender-affirming hormone treatment - A unique approach to study the effects of sex hormones on brain structure and function. Cortex 129:68–79, DOI: 10.1016/j.cortex.2020.04.005.

Leranth C, Hajszan T, MacLusky NJ (2004) Androgens increase spine synapse density in the CA1 hippocampal subfield of ovariectomized female rats. J Neurosci 24:495–499, DOI: 10.1523/JNEUROSCI.4516-03.2004.

Leranth C, Petnehazy O, MacLusky NJ (2003) Gonadal hormones affect spine synaptic density in the CA1 hippocampal subfield of male rats. J Neurosci 23:1588–1592, DOI.

MacLusky NJ, Hajszan T, Johansen JA, Jordan CL, Leranth C (2006) Androgen effects on hippocampal CA1 spine synapse numbers are retained in Tfm male rats with defective androgen receptors. Endocrinology 147:2392–2398, DOI: 10.1210/en.2005-0673.

Maudsley AA, Domenig C, Govind V, Darkazanli A, Studholme C, Arheart K, Bloomer C (2009) Mapping of brain metabolite distributions by volumetric proton MR spectroscopic imaging (MRSI). Magn Reson Med 61:548–559, DOI: 10.1002/mrm.21875.

McCarthy MM, Auger AP, Perrot-Sinal TS (2002) Getting excited about GABA and sex differences in the brain. Trends in neurosciences 25:307–312, DOI: 10.1016/s0166-2236(02)02182-3.

McEwen BS, Akama KT, Spencer-Segal JL, Milner TA, Waters EM (2012) Estrogen effects on the brain: actions beyond the hypothalamus via novel mechanisms. Behavioral neuroscience 126:4–16, DOI: 10.1037/a0026708.

McEwen BS, Milner TA (2017) Understanding the broad influence of sex hormones and sex differences in the brain. Journal of neuroscience research 95:24–39, DOI: 10.1002/jnr.23809.

Milner TA, McEwen BS, Hayashi S, Li CJ, Reagan LP, Alves SE (2001) Ultrastructural evidence that hippocampal alpha estrogen receptors are located at extranuclear sites. The Journal of comparative neurology 429:355–371, DOI.

Nagae-Poetscher LM, Bonekamp D, Barker PB, Brant LJ, Kaufmann WE, Horska A (2004) Asymmetry and gender effect in functionally lateralized cortical regions: a proton MRS imaging study. J Magn Reson Imaging 19:27–33, DOI: 10.1002/jmri.10429.

O’Gorman RL, Michels L, Edden RA, Murdoch JB, Martin E (2011) In vivo detection of GABA and glutamate with MEGA-PRESS: reproducibility and gender effects. J Magn Reson Imaging 33:1262–1267, DOI: 10.1002/jmri.22520.

Ohba S, Ikeda T, Ikegaya Y, Nishiyama N, Matsuki N, Yamada MK (2005) BDNF locally potentiates GABAergic presynaptic machineries: target-selective circuit inhibition. Cereb Cortex 15:291–298, DOI: 10.1093/cercor/bhh130.

Pandya M, Palpagama TH, Turner C, Waldvogel HJ, Faull RL, Kwakowsky A (2019) Sex-and age-related changes in GABA signaling components in the human cortex. Biology of sex differences 10:5, DOI: 10.1186/s13293-018-0214-6.

Peterson CM, Matthews A, Copps-Smith E, Conard LA (2017) Suicidality, Self-Harm, and Body Dissatisfaction in Transgender Adolescents and Emerging Adults with Gender Dysphoria. Suicide & life-threatening behavior 47:475–482, DOI: 10.1111/sltb.12289.

Pletzer B, Harris T, Hidalgo-Lopez E (2018) Subcortical structural changes along the menstrual cycle: beyond the hippocampus. Sci Rep 8:16042, DOI: 10.1038/s41598-018-34247-4.

Poeppl TB, Langguth B, Rupprecht R, Safron A, Bzdok D, Laird AR, Eickhoff SB (2016) The neural basis of sex differences in sexual behavior: A quantitative meta-analysis. Frontiers in neuroendocrinology 43:28–43, DOI: 10.1016/j.yfrne.2016.10.001.

Rai AL, Jeswar U (2012) Immunohistochemical colocalization of estrogen receptor-alpha and GABA in adult female rat hippocampus. Annals of neurosciences 19:112–115, DOI: 10.5214/ans.0972.7531.190305.

Reddy DS (2004) Testosterone modulation of seizure susceptibility is mediated by neurosteroids 3alpha-androstanediol and 17beta-estradiol. Neuroscience 129:195–207, DOI: 10.1016/j.neuroscience.2004.08.002.

Ruigrok AN, Salimi-Khorshidi G, Lai MC, Baron-Cohen S, Lombardo MV, Tait RJ, Suckling J (2014) A meta-analysis of sex differences in human brain structure. Neurosci Biobehav Rev 39:34–50, DOI: 10.1016/j.neubiorev.2013.12.004.

Saleh MG, Near J, Alhamud A, van der Kouwe AJW, Meintjes EM (2017) Effects of tissue and gender on macromolecule suppressed gamma-aminobutyric acid. Int J Imaging Syst Technol 27:144–152, DOI: 10.1002/ima.22218.

Sanacora G, Mason GF, Rothman DL, Behar KL, Hyder F, Petroff OA, Berman RM, Charney DS, Krystal JH (1999) Reduced cortical gamma-aminobutyric acid levels in depressed patients determined by proton magnetic resonance spectroscopy. Arch Gen Psychiatry 56:1043–1047, DOI.

Sanacora G, Treccani G, Popoli M (2012) Towards a glutamate hypothesis of depression: an emerging frontier of neuropsychopharmacology for mood disorders. Neuropharmacology 62:63–77, DOI: 10.1016/j.neuropharm.2011.07.036.

Scharfman HE, MacLusky NJ (2014) Differential regulation of BDNF, synaptic plasticity and sprouting in the hippocampal mossy fiber pathway of male and female rats. Neuropharmacology 76 Pt C:696–708, DOI: 10.1016/j.neuropharm.2013.04.029.

Seiger R, Hahn A, Hummer A, Kranz GS, Ganger S, Woletz M, Kraus C, Sladky R, Kautzky A, Kasper S, Windischberger C, Lanzenberger R (2016) Subcortical gray matter changes in transgender subjects after long-term cross-sex hormone administration. Psychoneuroendocrinology 74:371–379, DOI: 10.1016/j.psyneuen.2016.09.028.

Selakovic D, Joksimovic J, Jovicic N, Mitrovic S, Mihailovic V, Katanic J, Milovanovic D, Pantovic S, Mijailovic N, Rosic G (2019) The Impact of Hippocampal Sex Hormones Receptors in Modulation of Depressive-Like Behavior Following Chronic Anabolic Androgenic Steroids and Exercise Protocols in Rats. Frontiers in behavioral neuroscience 13:19, DOI: 10.3389/fnbeh.2019.00019.

Shannon M, Wilson J, Xie Y, Connolly L (2019) In vitro bioassay investigations of suspected obesogen monosodium glutamate at the level of nuclear receptor binding and steroidogenesis. Toxicology letters 301:11–16, DOI: 10.1016/j.toxlet.2018.10.021.

Silberbauer LR, Spurny B, Handschuh P, Klobl M, Bednarik P, Reiter B, Ritter V, Trost P, Konadu ME, Windpassinger M, Stimpfl T, Bogner W, Lanzenberger R, Spies M (2020) Effect of Ketamine on Limbic GABA and Glutamate: A Human In Vivo Multivoxel Magnetic Resonance Spectroscopy Study. Frontiers in psychiatry 11:549903, DOI: 10.3389/fpsyt.2020.549903.

Skilbeck KJ, Johnston GA, Hinton T (2010) Stress and GABA receptors. Journal of neurochemistry 112:1115–1130, DOI: 10.1111/j.1471-4159.2009.06539.x.

Spizzirri G, Duran FLS, Chaim-Avancini TM, Serpa MH, Cavallet M, Pereira CMA, Santos PP, Squarzoni P, da Costa NA, Busatto GF, Abdo CHN (2018) Grey and white matter volumes either in treatment-naive or hormone-treated transgender women: a voxel-based morphometry study. Sci Rep 8:736, DOI: 10.1038/s41598-017-17563-z.

Spurny B, Heckova E, Seiger R, Moser P, Klobl M, Vanicek T, Spies M, Bogner W, Lanzenberger R (2019) Automated ROI-Based Labeling for Multi-Voxel Magnetic Resonance Spectroscopy Data Using FreeSurfer. Frontiers in molecular neuroscience 12:28, DOI: 10.3389/fnmol.2019.00028.

Spurny B, Vanicek T, Seiger R, Reed MB, Klobl M, Ritter V, Unterholzner J, Godbersen GM, Silberbauer LR, Pacher D, Klug S, Konadu ME, Gryglewski G, Trattnig S, Bogner W, Lanzenberger R (2021) Effects of SSRI treatment on GABA and glutamate levels in an associative relearning paradigm. Neuroimage 117913, DOI: 10.1016/j.neuroimage.2021.117913.

Syan SK, Minuzzi L, Costescu D, Smith M, Allega OR, Coote M, Hall GBC, Frey BN (2017) Influence of endogenous estradiol, progesterone, allopregnanolone, and dehydroepiandrosterone sulfate on brain resting state functional connectivity across the menstrual cycle. Fertil Steril 107:1246–1255 e1244, DOI: 10.1016/j.fertnstert.2017.03.021.

Tabori NE, Stewart LS, Znamensky V, Romeo RD, Alves SE, McEwen BS, Milner TA (2005) Ultrastructural evidence that androgen receptors are located at extranuclear sites in the rat hippocampal formation. Neuroscience 130:151–163, DOI: 10.1016/j.neuroscience.2004.08.048.

Tsakiris M, Hesse MD, Boy C, Haggard P, Fink GR (2007) Neural signatures of body ownership: a sensory network for bodily self-consciousness. Cereb Cortex 17:2235–2244, DOI: 10.1093/cercor/bhl131.

Wilkinson ID, Paley MN, Miszkiel KA, Hall-Craggs MA, Kendall BE, Chinn RJ, Harrison MJ (1997) Cerebral volumes and spectroscopic proton metabolites on MR: is sex important? Magn Reson Imaging 15:243–248, DOI: 10.1016/s0730-725x(96)00334-7.

Yague JG, Azcoitia I, DeFelipe J, Garcia-Segura LM, Munoz A (2010) Aromatase expression in the normal and epileptic human hippocampus. Brain Res 1315:41–52, DOI: 10.1016/j.brainres.2009.09.111.

Zanos P, Gould TD (2018) Mechanisms of ketamine action as an antidepressant. Mol Psychiatry 23:801–811, DOI: 10.1038/mp.2017.255.

Zanos P, Moaddel R, Morris PJ, Georgiou P, Fischell J, Elmer GI, Alkondon M, Yuan P, Pribut HJ, Singh NS, Dossou KS, Fang Y, Huang XP, Mayo CL, Wainer IW, Albuquerque EX, Thompson SM, Thomas CJ, Zarate CA, Jr., Gould TD (2016) NMDAR inhibition-independent antidepressant actions of ketamine metabolites. Nature 533:481–486, DOI: 10.1038/nature17998.

