## Supplement material for "Effects of sex hormones on brain GABA and glutamate levels in a cis- and transgender cohort"

**Supplement table 1:** Treatment regimes in TM subjects

| treatment | number of subjects |
|---|---|
| Nebido 1000mg, every 8-12 weeks, i.m | 5 |
| testosterone cream 50mg/d | 4 |
| Nebido 1000mg + Moniq Gynial 75µg/d (desogestrel gel) | 2 |
| Testavan 46mg/d (testosterone gel) | 1 |
| testosterone cream 37.5mg/d + Moniq Gynial 75µg/d | 1 |
| testosterone cream 50mg/d + Moniq Gynial 75µg/d | 1 |
| Nebido 1000mg + decapeptyl depot (triptorelin), every 4-5 weeks | 1 |

**Supplement table 2:** Absolute number of data points (of max. 15) at each measurement included in linear mixed effect model analyses for each region and neurotransmitter ratio

| | | GABA+/tCr | | Glx/tCr | |
|---|---|---|---|---|---|
| | | **M1** | **M2** | **M1** | **M2** |
| **hippocampus** | CW | 15 | 12 | 15 | 15 |
| | TM | 13 | 13 | 13 | 14 |
| **insula** | CW | 12 | 13 | 14 | 15 |
| | TM | 12 | 12 | 14 | 15 |
| **putamen** | CW | 14 | 13 | 15 | 15 |
| | TM | 12 | 12 | 15 | 14 |
| **pallidum** | CW | 15 | 13 | 15 | 15 |
| | TM | 12 | 12 | 15 | 14 |
| **thalamus** | CW | 15 | 13 | 15 | 15 |
| | TM | 12 | 12 | 14 | 15 |

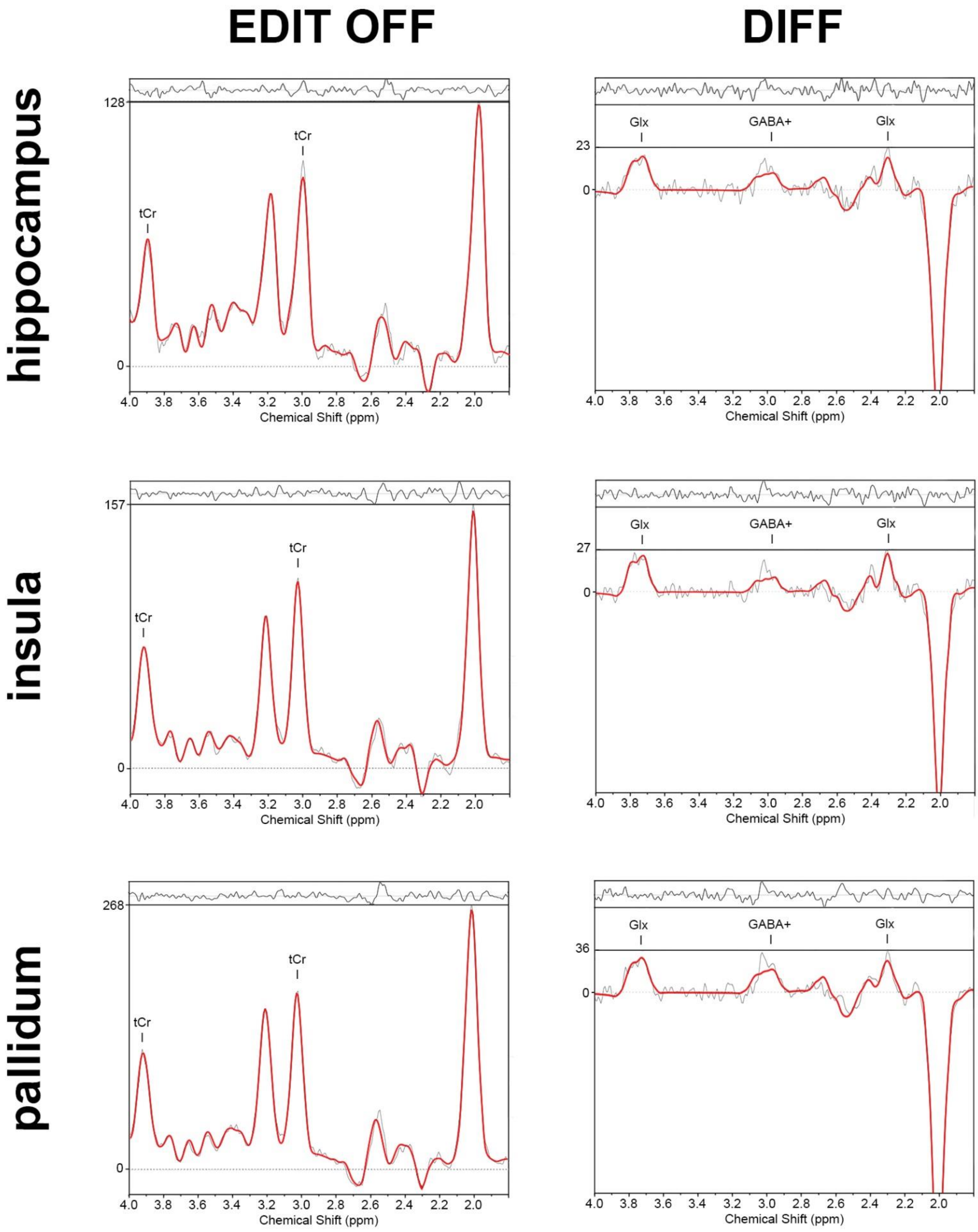


**Supplement figure 1:** LCModel fits of exemplary non-edited (EDIT OFF) and difference spectra (DIFF) of the hippocampus, insula and pallidum.
